# Scaffolding the Attention-Deficit/Hyperactivity Disorder Brain Using Random Noise Stimulation

**DOI:** 10.1101/19005983

**Authors:** Itai Berger, Ornella Dakwar-Kawar, Ephraim S. Grossman, Mor Nahum, Roi Cohen Kadosh

**Author notes:** Correspondence to: Dr. Itai Berger, MD, The Pediatric Neurology Service, Pediatric Division, Assuta Ashdod University Hospital, Faculty of Health Sciences, Ben-Gurion, University of the Negev, 7 Harefua St. Ashdod, Israel, Email address, Prof. Roi Cohen Kadosh, PhD, Department of Experimental Psychology, University of Oxford, New Radcliffe House, Oxford, OX2 6GG, England. Equally contributed (Itai Berger, Ornella Dakwar-Kawar).

## Abstract

Various methods have been attempted to effectively ameliorate psychiatric and neurological conditions in children and adults. One of the attractive ideas is to develop interventions to create a lasting, rather than only an immediate, effect. Neurostimulation has been shown to yield long-term effect when combined with cognitive training in healthy young adults. We examined whether such approach could benefit children with attention deficit hyperactivity disorder (ADHD), the most common neurodevelopmental disorder in childhood. We used a randomized double-blind active-controlled crossover study of 19 unmedicated children (aged 7–12 years old) with attention deficit hyperactivity disorder, who received either transcranial direct current stimulation or random noise stimulation while completing five-day executive functions training, which includes working memory, cognitive flexibility, and inhibition tasks. Both stimulation protocols have previously shown potential for inducing lasting benefits in adults, while transcranial direct current stimulation was examined in multiple attention deficit hyperactivity disorder studies and has been highlighted as a promising method for treating neuropsychological deficits. For our primary outcome, transcranial random noise stimulation yielded a clinical improvement as indicated by the reduced attention deficit hyperactivity disorder rating scale score from baseline, and in comparison to the changes observed in transcranial direct current stimulation. Moreover, the effect of brain stimulation one week after completion of treatment yielded further improvement, suggesting a neuroplasticity-related effect. Finally, transcranial random noise stimulation improved working memory compared to transcranial direct current stimulation, and a larger transcranial random noise stimulation effect on attention deficit hyperactivity disorder rating scale was predicted for those patients who showed the greatest improvement in working memory. Our results provide a promising direction toward a novel intervention in attention deficit hyperactivity disorder, which is shown to have a lasting effect via the modulating of neuroplasticity, rather than a merely immediate effect as was shown for in previous medical interventions.

## Introduction

Attention deficit hyperactivity disorder (ADHD) is the most common neurodevelopmental disorder in childhood, with significant negative lifetime outcomes (Greydanus *et al*., 2007). Despite proven effects of combinations of pharmacological and psychosocial interventions, there is still a need for improvement of cognitive dysfunction and behavioral symptoms that are only partially covered by current interventions (Moldavsky and Sayal, 2013). These factors highlight the pressing need for novel, efficacious interventions.

Transcranial electrical stimulation (tES) has been suggested as a possible noninvasive means to modify brain activity and steadily enhance behavioral and cognitive performance (Santarnecchi *et al*., 2015; Fertonani and Miniussi, 2017; Polania *et al*., 2018; Reed and Cohen Kadosh, 2018). Based on promising outcomes, one form of tES (namely, trigeminal nerve stimulation) has recently received FDA approval as a treatment for children diagnosed with ADHD who are not currently taking prescription ADHD medication (Voelker, 2019).

tES involves the application of a weak current (mostly 1–2 mA) to the brain via skin-electrode interface, creating an electric field that modulates neuronal activity (Polania *et al*., 2018). In the present research we used two types of tES: transcranial direct current stimulation (tDCS) and transcranial random noise stimulation (tRNS) (Paulus, 2011; Santarnecchi *et al*., 2015; Antal and Herrmann, 2016). tDCS is the most frequently used form of tES (Santarnecchi *et al*., 2015; Polania *et al*., 2018), and is used to modulate neuronal excitability in a subtle manner without depolarizing action potentials (Paulus, 2011; Antal and Herrmann, 2016; Fertonani and Miniussi, 2017). It has been suggested that the cortex beneath the anodal electrode typically becomes more excitable whereas the cathodal site has decreased excitability (Truong *et al*., 2014; Frangou *et al*., 2018). The delivery of tRNS uses the same equipment as tDCS to stimulate neuronal activity at intensities that do not lead to action potentials. However, the mechanisms by which tRNS influences brain activity are different (Chaieb *et al*., 2015; Fertonani and Miniussi, 2017). For example, for tDCS N-methyl-D-aspartate receptors have been shown to play a key role, with both the short and long-term effects of tDCS not being observed after blocking Na^+^ channels or after the administration of an N-methyl-D-aspartate receptor antagonist (Liebetanz *et al*., 2002; Nitsche *et al*., 2003). In contrast, the excitability enhancing effects of tRNS are significantly decreased by blocking voltage gated sodium channels, and the effect is likely to be independent of N-methyl-D-aspartate receptors (Chaieb *et al*., 2015).

In addition, in tRNS both electrodes can be used to increase cortical excitability, either in homologous locations bilaterally or at different regions simultaneously (Terney *et al*., 2008; Snowball *et al*., 2013). Previous studies, mainly in healthy young adults, have shown that when several sessions of tDCS or tRNS are applied during cognitive training, the effects can last from weeks to months (Reis *et al*., 2009; Baker *et al*., 2010; Cappelletti *et al*., 2013; Snowball *et al*., 2013; Looi *et al*., 2016; Frank *et al*., 2018; Brevet-Aeby *et al*., 2019). In children and adolescents, who might show accelerated neural plasticity compared to adults (for a review see Cramer *et al*., 2011), tES combined with behavioral intervention has been suggested as a useful tool to modulate neuroplasticity in those with atypical development to generate long-lasting effects (Brunoni *et al*., 2012; Krause and Cohen Kadosh, 2013; Costanzo *et al*., 2016; Costanzo *et al*., 2019). The excellent safety profile of tES makes it an even more appealing treatment method for children and adolescents (Krishnan *et al*., 2015; Antal *et al*., 2017).

One of the most influential theories of the neural basis of ADHD suggests that deficient inhibitory control mechanisms give rise to executive dysfunction, which is likely genetically influenced (Sonuga-Barke, 2005; Goos *et al*., 2009). The neuroanatomic substrate of inhibitory control has been imputed to the basal ganglia-thalamo-cortical circuits (Aron *et al*., 2004; Christakou *et al*., 2004; Norman *et al*., 2016). Specifically, this network links the prefrontal cortex to the dorsal neo-striatum via excitatory glutaminergic cells, the basal ganglia to the dorsomedial thalamus via inhibitory projections, and the thalamus back to the prefrontal cortex via excitatory projections (Castellanos *et al*., 2002). Inhibitory control is processed during the maturation of this circuit. Previous studies have shown that ADHD is associated with structural and functional abnormalities within this circuit (Castellanos *et al*., 2002). Indeed, several studies have successfully used anodal tDCS over the left dorsolateral prefrontal cortex (dlPFC) in children with ADHD (Munz *et al*., 2015; Bandeira *et al*., 2016; Nejati *et al*., 2017; Allenby *et al*., 2018; Soltaninejad *et al*., 2019) (for reviews see (Rubio *et al*., 2015; Palm *et al*., 2016)). An updated meta-analysis suggests that anodal tDCS over the left dlPFC can yield a small-to-medium effect size (a cumulative effect size of (Ē)=0.255– 0.681) on neuropsychological deficits, such as inhibition and working memory, in ADHD (Salehinejad *et al*., 2019).

To the best of our knowledge, tRNS has not yet been used in the case of ADHD, as it is a more novel form of brain stimulation (Santarnecchi *et al*., 2015; Antal and Herrmann, 2016). However, tRNS in healthy adults successfully improved high-level cognitive functions including attentional control, with stronger effects shown for individuals with a phenotype that indicates poorer attentional control (Harty and Kadosh, 2019). In a small sample (n=6 per group, between-subject design) of atypically developing children with dyscalculia, 0.75mA tRNS over bilateral dlPFC during numerical training has shown positive effects on numerical training compared to sham (placebo) stimulation (Looi *et al*., 2017). This preliminary result is in line with the concept that random noise can have beneficial effects on behavior, and it is supported by earlier animal research that suggest that random noise can allow greater opportunity for neuroplasticity (Chang and Merzenich, 2003).

The goal of our study was to compare the beneficial effects of tRNS and tDCS when combined with executive function (EF) training in ameliorating symptoms and EF in unmedicated school-age children with ADHD. The computerized EF training (ACTIVATE(tm)) we utilised in the present study was composed of four different games, collectively targeting the core EF components of visual-spatial working memory, cognitive flexibility and inhibitory control (Diamond, 2013). We conducted a double-blind randomized controlled trial with a crossover design, in which each tES method was applied for 5 consecutive days along with EF training. We limited each arm to 5 days as previous tDCS and tRNS studies on healthy adults have yielded lasting effects with protocols of a similar duration or even shorter (Reis *et al*., 2009; Snowball *et al*., 2013; Looi *et al*., 2016; Pasqualotto, 2016; Frank *et al*., 2018). Endurance of effects was measured one week after the end of the intervention protocol. We used tDCS with a montage that has been deemed to be the most successful so far based on a meta-analysis of tDCS studies in ADHD, i.e., an anodal electrode over the left dlPFC and a cathodal electrode over the contralateral supraorbital (Salehinejad *et al*., 2019). With tRNS, we used a montage that placed the electrodes over the left dlPFC and the right IFG. This montage was chosen due both to the advantage of this neurostimulation polarity-independent method and to its ability to yield excitatory stimulation without parallel inhibitory effects (Terney *et al*., 2008). Moreover, a similar montage was used in previous tRNS studies in the field of cognitive training in healthy young adults and children with dyscalculia (Snowball *et al*., 2013; Popescu *et al*., 2016; Looi *et al*., 2017; Brem *et al*., 2018; Frank *et al*., 2018). If tES combined with EF training is effective in alleviating ADHD symptoms, it may offer many advantages as a relatively inexpensive, noninvasive therapeutic option for school-age children with ADHD.

## Methods and Materials

### Study Design

We conducted a randomized double-blind active-controlled crossover study of children diagnosed with ADHD. Twenty-two children were assessed for eligibility, 21 children were recruited for the study, and 19 participants completed it (see **Figure 1** for the CONSORT flow diagram). Two participants were excluded from the study: one of them due to complaints of an uncomfortable topical sensation and headaches during the tDCS protocol. The second participant was excluded as the parents reported in the third session behavior that might meet one of the exclusion criteria (the expression of self-harm thoughts), which was present already two months before study participation but was not reported at screening.

**Figure 1.**
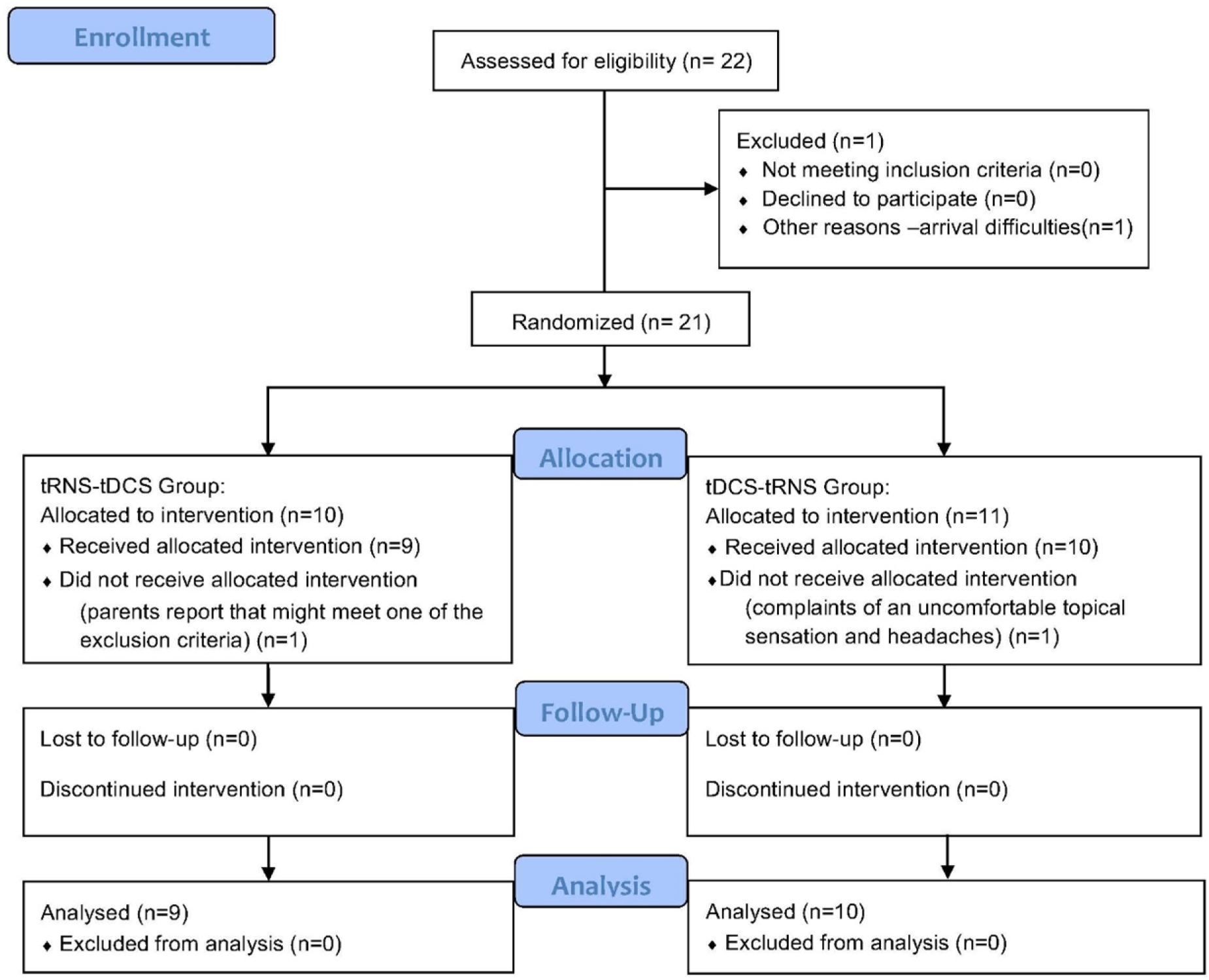
CONSORT flow diagram of the progress through the phases of the randomized crossover study of the two groups.

The study design scheme is shown in **Figure 2**. Following screening, eligible participants were assessed at baseline and then randomized into receiving either tDCS or tRNS first in week 1, along with computerized EF training. Each group received either tDCS or tRNS treatment for 5 consecutive days (one treatment session each day). Following a one-week break, there was a crossover between the groups in week 3: those who received tDCS in the first week received tRNS in the third week, while those who received tRNS in the first week received tDCS in the third week. This allowed us to compare the different treatment in a within-subject design, as well as to examine one-week post-treatment effects to assess lasting effects. The assessment battery was repeated at the end of each week. The total duration of subject participation in the study was 4 weeks. All study-related activities were conducted in a research lab at the School of Occupational Therapy of the Hebrew University of Jerusalem.

**Figure 2.**
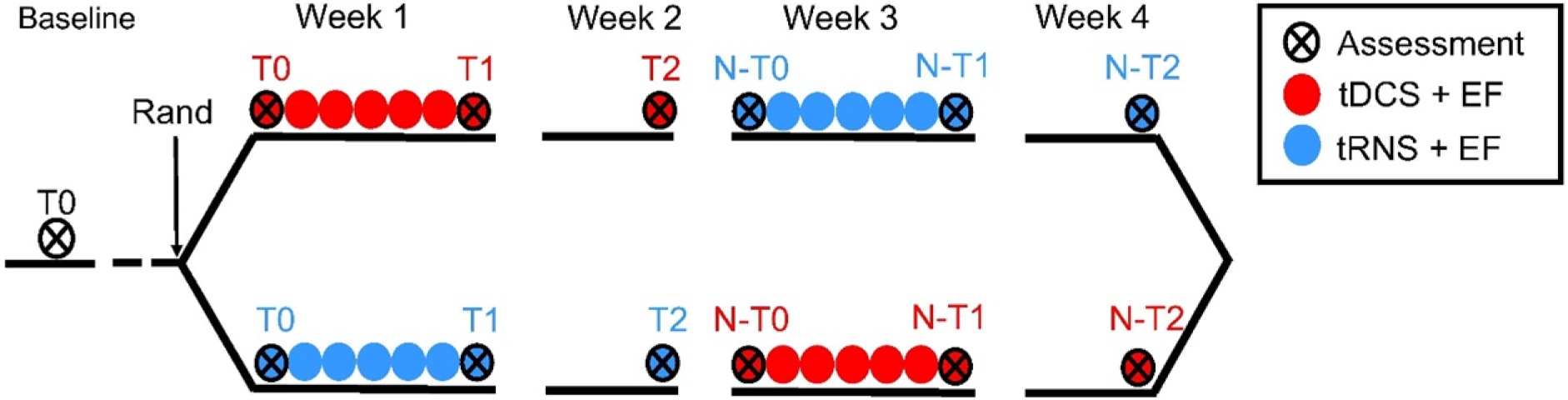
Study Design. Eligible participants with ADHD were randomized into one of two treatment groups. Baseline measures were acquired (T0) before the randomization into one of the two groups. Participants in both groups received 5 daily treatment sessions in Week 1 and were assessed at the end of this week (T1). At Week 2 no treatment was given and the lasting effect from week 1 was measured (T2). On Week 3 the participants received the treatment that the other group received in Week 1. That is, if participants received tRNS+EF on Week 1, they received tDCS+EF on Week 3. To allow an accurate assessment of the new treatment on Week 3, we recalibrated the participants’ baseline measures by using their latest assessment data from T2. This new baseline measure was called New T0 (N-T0). We reassessed the participants at the end of Week 3 (N-T1). At Week 4 no treatment was given and the lasting effect from week 3 was measured (N-T2).

### Study Population

The study included children aged 7–12 years old. Participants were recruited among children referred to the ADHD clinic by pediatricians, general practitioners, teachers, psychologists, or parents. All participants agreed to participate in the study (verbal assent) and their parents gave written informed consent to the study, approved by the Helsinki Committee (IRB) of the Hebrew University and Hadassah Medical Center (Jerusalem, Israel).

The local IRB approved a total number of 100 participants for this study. For safety reasons we were asked to summarise the data of the first 20 participants in order to assess safety and tolerability. Upon clinical review by the study team and the IRB - if all safety criteria are met, the study will proceed to recruit another 80 participants. That is the reason we chose to include all 20 participants in the tDCS-tRNS arm, so safety and tolerability, as well as efficacy, will be assessed for both methods and will allow us to revise accordingly the testing plans for the future participants. The study is registered at ClinicalTrials.gov (identifier NCT03104972).

A power analysis revealed that the obtained sample size, power=.8, and α=.05 would allow to detect an effect with an obtained effect size of .68. This is due to the within-subject design, which allows, with the given sample size, for more power to detect an effect compared to previous studies, including a recent trial that used a between-subject design (n=32 in one group and n=30 in another) and led to FDA approval of its use in brain stimulation to treat ADHD (McGough *et al*., 2019; Voelker, 2019).

#### Inclusion criteria

Each child met the criteria for ADHD according to DSM–5 (American Psychiatric Association, 2013), using the “gold standard” procedure as described by the American Academy of Pediatrics, and including a semi-structured interview of the patient and parents by a specialist in pediatric neurology and child development, a neurological examination, and ADHD rating scale (ADHD-RS) diagnostic questionnaires (DuPaul *et al*., 1998; DuPaul *et al*., 2016). Each child scored above the standard clinical cutoff values for ADHD symptoms on ADHD DSM–5 scales (DuPaul *et al*., 1998; American Psychiatric Association, 2013; DuPaul *et al*., 2016). All children were newly diagnosed and drug naïve.

#### Exclusion criteria

Children were excluded from the study if they had one of the following: a chronic neurological disease, epilepsy in the participant or in a first-degree relative, intellectual disability, other chronic conditions, chronic use of medications, or other primary psychiatric diagnosis (e.g., depression, anxiety, psychosis). The Hebrew translation of the Kiddie-SADS-Lifetime Version (Kaufman *et al*., 2000) was used to assess axis-I disorders in participants according to DSM–5 criteria (Kaufman *et al*., 2000).

Prospective resting-state electroencephalography was performed at screening in order to rule out an unknown existence of epileptiform activity. Electroencephalography records were standardized and recorded with gTech’s g.Recorder software, using a 64-channel wireless electroencephalography cap system (g.Nautilus) with gel-based electrodes.

### Primary Outcome Measure

The primary outcome measure of the study is the total score of the ADHD-RS diagnostic questionnaire completed by the parents (DuPaul *et al*., 1998; DuPaul *et al*., 2016). This scale is of well-accepted validity and reliability, regarded as standards in ADHD diagnosis and treatment effect (Snowball *et al*., 2013). The ADHD-RS-5 contains 18 items based on the wording used to describe those items in the DSM–5. The 18 items are presented in the context of a two-factor structure beginning with the nine inattention (IN) symptoms followed by the nine hyperactive-impulsive (HI) symptoms. Parents rate each of these items on a 4-point Likert frequency scale that can be scored 0 (never or rarely), 1 (sometimes), 2 (often), or 3 (very often). IN and HI total symptom severity scores categorically generate IN and HI symptom counts. The symptom count for IN is determined by summing the number of IN items receiving ratings of 2 (often) or 3 (very often). The symptom count for HI is calculated in a similar fashion. Thus, for both IN and HI, symptom counts range from 0 to 9 in accordance with DSM–5 criteria and 18 is the maximal possible scoring for the entire scale (Anastopoulos *et al*., 2018).

### Secondary outcome measures

1. <u>CGI-S</u> (Clinical Global Impression – Severity) <u>scale</u>: a 3-item observer-rated scale that measures illness severity, as assessed by the treating clinician (Guy, 1976). Scoring the CGI-S is rated on a 7-point scale, with the severity of illness scale ranging from 1 (normal) to 7 (severely ill).
2. <u>MOXO-CPT</u> (NeuroTech Solutions Ltd): a standardized computerized test that measures attentional performance (Berger *et al*., 2017). The MOXO-CPT includes four performance indices: attention, timing, impulsivity, and hyperactivity.
3. <u>Digit Span:</u> a subtest of the Wechsler Intelligence Scale for Children – Fourth edition that measures short-term auditory memory and attention (Wechsler, 2003).

### Study Interventions

Participants completed computerized EF training along with either tDCS or tRNS brain stimulation.

#### Computerized EF Training

Participants completed training using the ACTIVATE^™^ training program, delivered on a tablet (Wexler *et al*., 2016). This gamified EF training includes different mini-games that target different EF components: working memory, cognitive flexibility, response inhibition, and sustained attention (Wexler *et al*., 2016). Each training session included 4 mini-games, each played for 5 minutes, for a total duration of 20 minutes of gameplay per session, which coincided with the tES protocol. The training starts at a basic level and adaptively progresses to move advanced levels, which include more complex tasks, depending on individual performance. The inclusion of several mini-games within a single session decreases the likelihood of contextualization (which is often the result of extensive training using a single task) and increases the likelihood of transfer (Perlman *et al*., 2016). While the present cognitive training has been used in previous studies that aimed to improve academic performance in typically developing children (Wexler *et al*., 2016), and in some preliminary studies in children with ADHD (de Oliveira Rosa *et al*., 2019), the outcome measures in previous studies never included ADHD-RS, which prevented us from estimating the effect size expected from such training alone. As our study focused on comparing the efficacy of two tES methods, rather than on EF training per se, a detailed description of the EF training protocol is beyond the scope of this paper, but can be found elsewhere (see (Wexler *et al*., 2016)) and the supplementary information section.

#### Transcranial Electrical Stimulation

Both tDCS and tRNS were applied using semi-dry 5X5 cm electrodes using the NovoStim device (Tech InnoSphere Eng. Ltd., Haifa). The NovoStim device is a research and investigational device, pending FDA and medical CE approval. Stimulation was delivered for 20 minutes each session, while participants completed the cognitive training (**Figure 3**). The total stimulation time for each tES protocol was 100 minutes (5 sessions of 20 min each).

**Figure 3.**
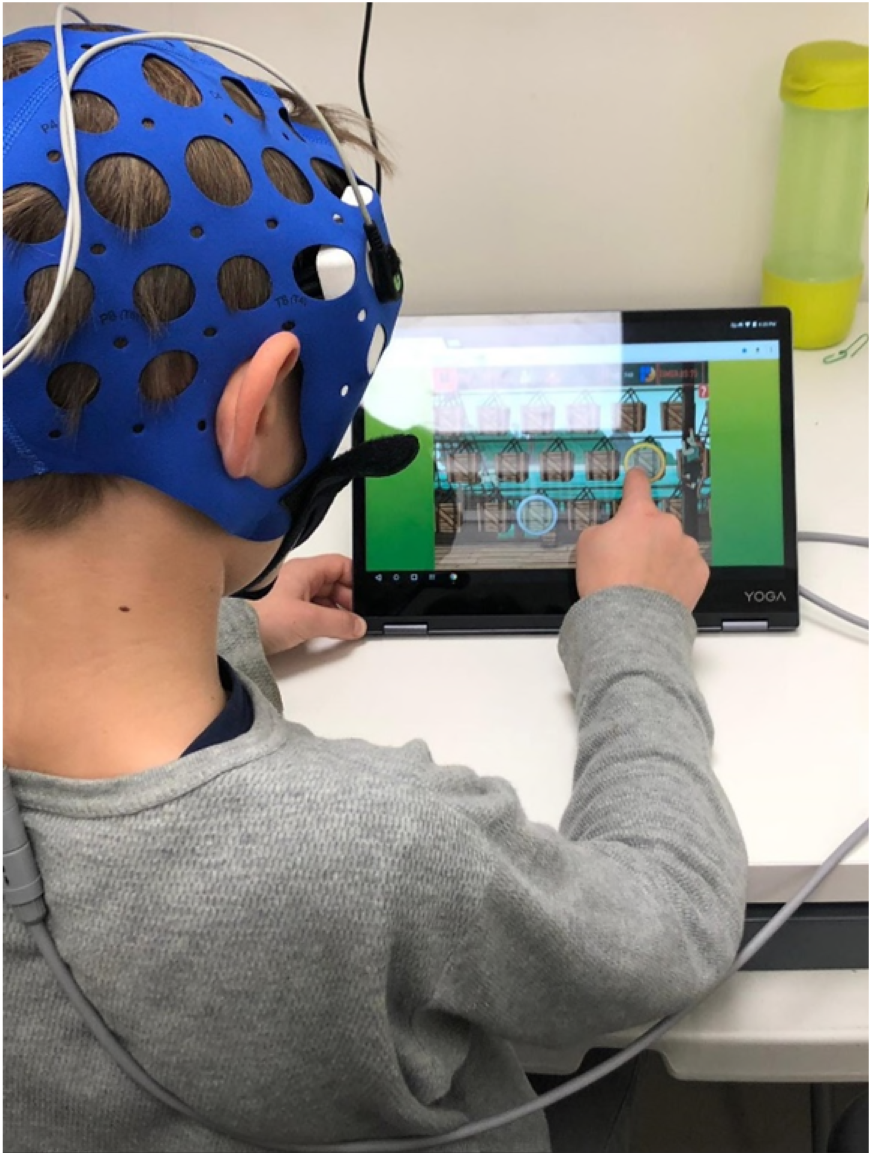
A treatment session of tES combined with EF training. Participants completed 20 minutes of EF training while tRNS or tDCS was delivered to them during this period.

<u>tDCS</u>. The current was set to 0.75mA based on previous computational modelling of tDCS in children and is estimated to equal that of approximately 1.5mA in adults (Kessler *et al*., 2013). Ramp-up and ramp-down durations were 30 seconds each. These durations were chosen after considering the parameters that would influence current distribution and density at the site of stimulation, such as thinner scalp, less cerebrospinal fluid, and smaller head size of the pediatric population (Kessler *et al*., 2013). A similar dosage of tDCS was well tolerated by the children and was not associated with adverse effects (Krishnan *et al*., 2015). The anodal electrode was positioned above the left dlPFC (F3 based on the International 10-20 system), while the cathodal electrode was placed over the right supraorbital (Fp2).

<u>tRNS</u>. Stimulation was applied at an amplitude of 0.75mA of tRNS over the left dlPFC and the right inferior frontal gyrus (IFG), attached under designated electrode positions (F3-F8 based on the International 10-20 system) of the tES cap. These stimulation locations were chosen based on their involvement in executive control and inhibition processes (Castellanos *et al*., 2002; Aron *et al*., 2004; Christakou *et al*., 2004). Ramp-up and ramp-down durations were the same as in the tDCS condition.

To mitigate the possibility that the research assistant will notice the differences in the montages between tDCS and tRNS, and would be biased toward a given montage, we alternated three naive research assistants throughout this study.

### Statistical Analysis

To examine treatment effects, we used linear mixed effects models, which account for within-subject correlations more optimally compared to ANOVA and automatically handle missing values, allowing maximum use of available data (Seltman, 2009). We used the R-package *nlme* (Pinheiro *et al*., 2017) to perform the linear mixed effects analysis with maximized log-likelihood on the outcome measures. We examined outcomes immediately post-treatment and one week later for each stimulation type, and included stimulation type (tDCS, tRNS) and time (immediately after treatment and one week post-treatment) as predictors. We included baseline performance as a covariate in our model, rather than use a subtraction score (i.e., post-treatment minus baseline). Including baseline performance as a covariate allows for a better adjustment for minor differences in the pre-treatment means. In contrast, subtraction score contains measurement error from both the baseline performance and the post-treatment score and is also negatively correlated with baseline performance because of the measurement error (Edwards, 2001; Jamieson, 2007). This approach has been employed by our group in recent publications (Looi *et al*., 2017; Clayton *et al*., 2019). As we used a within-subjects design, we used the baseline measures at T0 for the first arm, but to allow an accurate assessment of second arm, we recalibrated the participants’ baseline measures by using their latest assessment data from T2 as the new baseline for the second arm (Figure 2).

For all the measures we verified that the residuals were normally distributed using a q-q plot and the Shapiro–Wilk normality test. The only exception was the MOXO-CPT residuals, which were not normally distributed; we therefore applied the Tukey ladder of powers transformation, which is recommended in this case (Tukey, 1977).We also tested for the inclusion of an interaction term in our analysis. In our primary outcome, ADHD-RS, the interaction between stimulation type and time was not significant [β=.14, SE=.18, t(35)= .78, p=.44, 95% confidence intervals (CI) (-.21, .5)]. A model comparison showed no benefit from a more complex model, favoring the more parsimonious model, which included the main effects of stimulation and time (chi-squared test=.66, p=.41). We therefore report this parsimonious model also for the secondary outcome measures. However, as with the other measures, the inclusion of the interaction term between stimulation and time was not significant. We also explored the effect of order (tRNS first followed by tDCS, vice versa), but this variable was not significant [β=-.12, SE=.12, t(34)= -1.04, p=.3, 95% CI (-.37, .11)], and a model comparison preferred the simpler model that did not include this variable (chi-squared test=1.12, p=0.29).

### Data availability

Data is available upon reasonable request from the first author.

## Results

### Side Effects and Safety Issues

There were 61 records of side effects reported, none of which were considered clinically significant. **Table 1** summarizes these findings as a function of brain stimulation. As can be seen, tRNS yielded fewer reports of side effects. This finding is in line with the relevant literature on adults that highlights tRNS as a more comfortable neurostimulation method in comparison to tDCS (Ambrus *et al*., 2010; Fertonani *et al*., 2015).

**Table 1.**
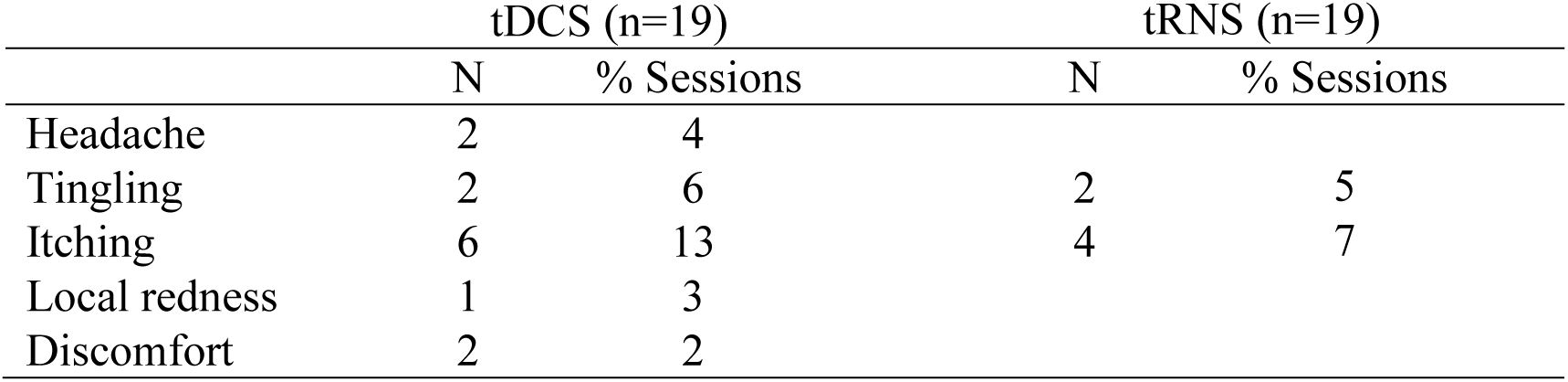
Spontaneously Reported or Observed Adverse Events during tDCS and tRNS. The table indicates the number of participants (N) and the percentage of sessions endorsing side effects at some point during the intervention.

|  | tDCS (n=19) |  | tRNS (n=19) |  |
| --- | --- | --- | --- | --- |
|  | N | % Sessions | N | % Sessions |
| Headache | 2 | 4 |  |  |
| Tingling | 2 | 6 | 2 | 5 |
| Itching | 6 | 13 | 4 | 7 |
| Local redness | 1 | 3 |  |  |
| Discomfort | 2 | 2 |  |  |

### Primary Outcome Measure: Changes in ADHD Symptoms

For the primary outcome, we predicted the ADHD-RS total score post-treatment immediately after the intervention (tRNS/tDCS) and one week later, while covarying for the baseline score. The analysis revealed a main effect of stimulation type, indicating greater improvement for tRNS than for tDCS [β=-.42 (SE=.18), B=-1.98 (SE=.87), t(35)=-2.28, p=.028, 95% CI (-3.67, -.29)] **Table 2**). The main effect of time, i.e., immediately after the end of the intervention to one week later, showed a further improvement one week after the end of the treatment [β=-.19 (SE=.09), B=-1.78 (SE=.86), t(35)=-2.07, p=.045), 95% CI (-3.46, -.1)]. In terms of improvement from baseline, tRNS yielded a mean improvement of 3.47 points [SE=1.03, t(15)=3.35, p=.004, 95% CI (1.31, 5.64)], while tDCS yielded a mean improvement of .57 points [SE=1.19, t(15)=.47, p=.64, 95% CI (-1.92, 3.06)].

**Table 2.**
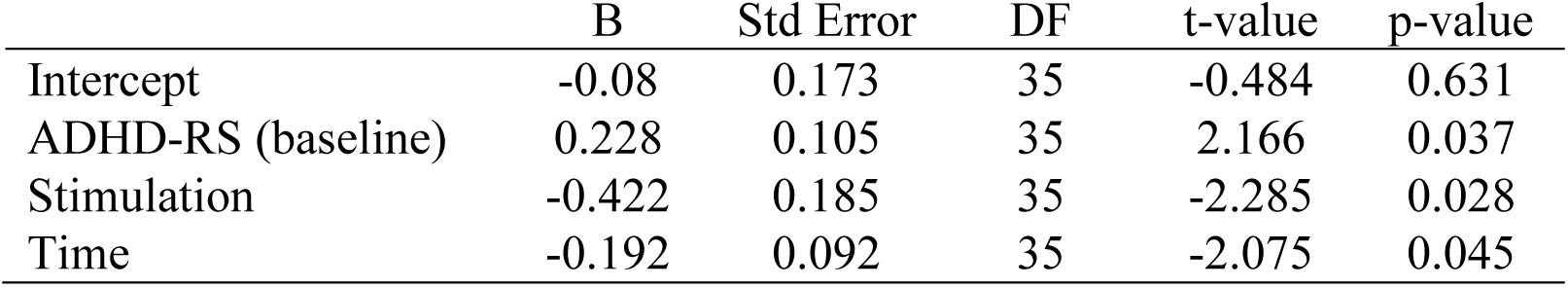
Beta Weights (Standardized) of the Regression Model with Post-treatment ADHD-RS Score as the Outcome Measure. The results indicate a significant effect for stimulation due to greater reduction in the ADHD-RS score for tRNS in comparison to tDCS, and greater improvement, as opposed to deterioration, as time passed following the treatment (one week later).

|  | B | Std Error | DF | t-value | p-value |
| --- | --- | --- | --- | --- | --- |
| Intercept | -0.08 | 0.173 | 35 | -0.484 | 0.631 |
| ADHD-RS (baseline) | 0.228 | 0.105 | 35 | 2.166 | 0.037 |
| Stimulation | -0.422 | 0.185 | 35 | -2.285 | 0.028 |
| Time | -0.192 | 0.092 | 35 | -2.075 | 0.045 |

### Changes in Secondary Outcome Measures

The secondary outcome measures were considered more exploratory. As such, we present them below without applying a correction for multiple comparisons, yet highlight that none of the results was significant at a α≤.05 after applying Bonferroni correction for multiple comparisons.

#### Changes in Attentional Performance

The results for the MOXO-CPT subscales and CGI-S (see **Tables S1–S5**) did not show a significant post-treatment effect of stimulation type (all ps>0.44), aside from the MOXO timing index, which showed larger changes following tRNS compared with those seen following tDCS [B=1.92 (SE=.91), t(47)=2.11, p=.04, 95% CI (.14, 3.7)]. However, as a transformation was applied to the outcome score due to the distribution of the residuals (see the Methods section), this result should be interpreted with caution, although the effect of stimulation was significant also without data transformation [β=.25 (SE=.12), B=.52 (SE=.25), t(47)=2.07, p=.044), 95% CI (.03, 1.02)].

#### Changes in Working Memory and in Short-Term Memory

Performance on the digit span subscale of the WISC (total score of forward and backward span) after the intervention showed similar results to those of the primary outcome (ADHD-RS), with a significant effect of stimulation, favoring tRNS over tDCS [β=.34 (SE=.14), B=1.07 (SE=.44), t(50)=2.44, p=.018, 95% CI (.22, 1.92)]. While the effect of time was not significant, descriptively it showed a positive, rather than a negative, slope, indicating that a deterioration due to the time elapsed from the end of the intervention did not occur [β=.04 (SE=.068), B=.27 (SE=.43), t(50)=.64, p=.53, 95% CI (-.56, 1.11)].

We further examined whether the improvement in the post-treatment digit span test reflects a modification in working memory or in short-term memory, as assessed by a backward and a forward digit span, respectively. Our results indicate that tRNS led to a significantly better performance in the backward digit span only, compared to tDCS [**Table 3**, backward digit span: β=.33 (SE=.16), B=.63 (SE=.3), t(51)=2.12, p=.038, 95% CI (.04, 1.22); forward digit span: β=.04 (SE=.16), B=.058 (SE=.24), t(51)=.24, p=.81, 95% CI (-.41, .52), **Table S6**].

**Table 3.** Beta Weights (Standardized) of the Regression Model with Post-treatment Backward Digit Span Score. The results indicate a significant effect for stimulation due to greater increase in the backward digit span score for tRNS in comparison to tDCS.

| | $\beta$ | Std Error | DF | t-value | p-value |
| --- | --- | --- | --- | --- | --- |
| Intercept | -0.197 | 0.108 | 51 | -1.811 | 0.076 |
| Stimulation | 0.331 | 0.156 | 51 | 2.123 | 0.038 |
| Backward digit span (baseline) | 0.84 | 0.077 | 51 | 10.928 | <0.001 |
| Time | 0.052 | 0.077 | 51 | 0.673 | 0.504 |

### Examining the Link between Clinical and Cognitive Changes by Brain Stimulation and Cognitive Training

Next, we examined whether the improvement in the ADHD-RS score under the tRNS protocol depends on the changes in WM performance (the backward digit span score). To do so, we ran a moderation analysis and predicted the post-treatment ADHD-RS score by stimulation type and the post-treatment backward digit span score, while controlling for the ADHD-RS and backward digit span scores at baseline. This analysis revealed a trend toward a significant interaction between stimulation type and the post-treatment backward digit span score [β=-.41 (SE=.23), B=-1.01 (SE=.56), t(53)=-1.81, p=.075, 95% CI (-2.09, 0.06), **Table 4**]. A simple slopes analysis revealed that this trend stemmed from a significant improvement in ADHD-RS for tRNS vs. tDCS in those who had showed the largest improvement in the backward digit span test [β=-.62 (SE=.29), B=-2.91 (SE=1.36), t(53)=-2.14, p=.037), 95% CI (-.24, -5.57)].

**Table 4.**
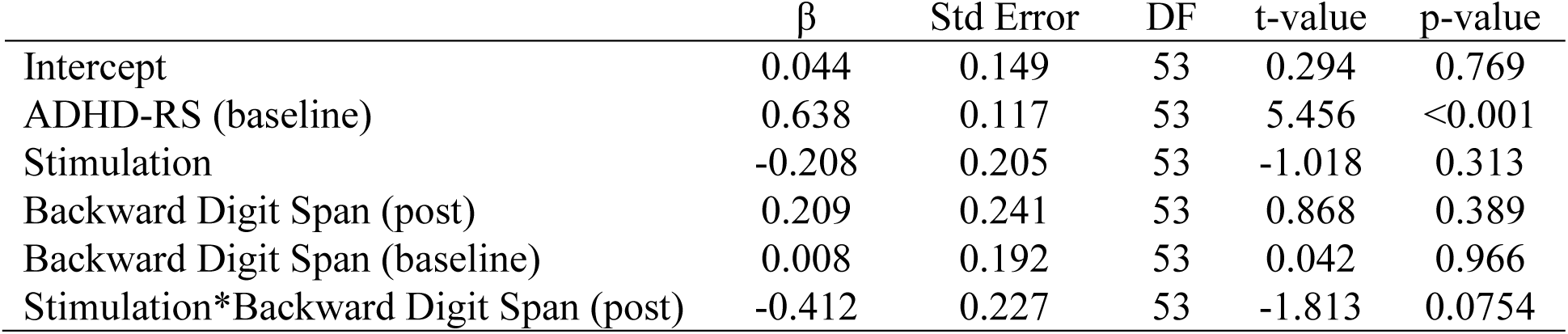
Moderation Analysis Predicting Post-treatment ADHD-RS Score. The results revealed a trend toward significant interaction between stimulation type and post-treatment (post) backward digit span in predicting the post-treatment ADHD-RS score.

| | $\beta$ | Std Error | DF | t-value | p-value |
| --- | --- | --- | --- | --- | --- |
| Intercept | 0.044 | 0.149 | 53 | 0.294 | 0.769 |
| ADHD-RS (baseline) | 0.638 | 0.117 | 53 | 5.456 | <0.001 |
| Stimulation | -0.208 | 0.205 | 53 | -1.018 | 0.313 |
| Backward Digit Span (post) | 0.209 | 0.241 | 53 | 0.868 | 0.389 |
| Backward Digit Span (baseline) | 0.008 | 0.192 | 53 | 0.042 | 0.966 |
| Stimulation*Backward Digit Span (post) | -0.412 | 0.227 | 53 | -1.813 | 0.0754 |

## Discussion

In this study we examined the effect of tRNS and tDCS on the clinical and cognitive outcomes of children with ADHD during 5 days of executive function training. The most notable results in our study are the improvements on ADHD-RS scores following tRNS and EF training relative to baseline and to tDCS and EF training. These promising results on the tRNS protocol support those of several studies in healthy young adults (Terney *et al*., 2008; Fertonani *et al*., 2011; Cappelletti *et al*., 2013; Snowball *et al*., 2013; Pasqualotto, 2016; Popescu *et al*., 2016; Brem *et al*., 2018; Frank *et al*., 2018; Brevet-Aeby *et al*., 2019; Peña *et al*., 2019), including a recent meta-analysis (Simonsmeier *et al*., 2018). Importantly, the results showed a further significant improvement 7 days after the end of the treatment, mirroring a similar lasting tRNS effect in previous studies on healthy adults (Cappelletti *et al*., 2013; Snowball *et al*., 2013; Pasqualotto, 2016; Frank *et al*., 2018; Brevet-Aeby *et al*., 2019). We suggest that the observed effects in the present study reflect our approach not to use brain stimulation alone, but to combine it with cognitive training in order to induce changes in the associated neural system via neuroplasticity (Cappelletti *et al*., 2013; Krause and Cohen Kadosh, 2013; Costanzo *et al*., 2019). This approach differs from other attempts to treat individuals with ADHD using drugs or brain stimulation alone.

The success of our approach is further supported by a tRNS effect on the MOXO timing index score, which reflects cognitive processing speed, i.e., the speed at which a person is able to perceive and react to stimuli in the environment (Nielsen *et al*., 2017). More importantly, we observed a tRNS effect on the backward digit span test, which measures WM capacity. This last effect was expected given that our cognitive training targeted WM as one of the executive functions that have been shown to be impaired in children with ADHD (Barkley, 1997). Notably, the effect of tRNS vs. tDCS was restricted to WM and did not extend to the forward digit span test, which does not measure WM but rather short-term memory span. Moreover, a greater improvement of ADHD-RS by tRNS was predicted by a greater improvement in the backward digit span test from baseline, while the interaction between brain stimulation and WM in predicting ADHD-RS was only marginally significant.

There are a few differences that are worth emphasizing when comparing our approach to that of that of McGough et al. (McGough *et al*., 2019), whose recent promising findings on trigeminal nerve stimulation as a treatment for ADHD has received FDA approval (Voelker, 2019). Our results are based on lower stimulation intensity (.75mA vs. 2–4mA) and shorter treatment duration (100min vs. 13,440min in total), and they show persistent and even increasing improvement after treatment, indicating plasticity-related effects. This is in contrast to the short-lived immediate improvement and significant deterioration one week after the end of the treatment associated with the form of tES reported in (McGough *et al*., 2019). Moreover, the estimated effect size in our study on ADHD-RS is higher than the one reported in (McGough *et al*., 2019) (estimated Cohen’s d=.95 on an 18 points scale, and .73 on a 54 points scale vs. .51). This difference is less likely to be due to an inflated effect size due to an underpowered design (Button *et al*., 2013) as the experimental design in our study was more suitable for detecting the observed effect size. While in our study we recruited 19 children (between May 2018 to March 2019), this sample size is the upper range of the sample size used in pediatric ADHD neurostimulation studies (with n=9-21, (Iadecola, 1993; Munz *et al*., 2015; Soltaninejad *et al*., 2015; Bandeira *et al*., 2016; Breitling *et al*., 2016; Soff *et al*., 2017; Sotnikova *et al*., 2017; Allenby *et al*., 2018)). Notably, most of these studies did not required the patients and their guardians to come to the lab multiple time as in the present study, which increase difficulties in terms of recruitment, and parental and child’s commitment. Moreover, all the children in our study were newly diagnosed and drug naïve.

Given the difficulties in recruiting such population, and for cognitive training which requires longer protocols, we chose in the present study to use a within-subject design. This approach allowed us to control better for individual differences that are impossible to perfectly match in a between-subject design, and at the same time allowing more powerful design, with the given sample size. For example, in a between-subject design the sample size required to detect an effect with α=.05, power (1-β)=.8, and an effect size of Cohen’s d=.68 using a t-test with a non-directional hypothesis is 72, more than 3.5 times the sample size required using a within-subject design.

However, one potential caveat is the lasting effect of a given intervention (e.g., tRNS). To take this into account we “rescaled” in our statistical model the baseline performance before the beginning of the second intervention, and also examine how the factor order, which was not significant, could influence the results. While we chose to examine the effect one week later, in order to reduce the attrition rate, and given the novelty of our approach, these results motivate future studies that will include a longer time period between the two treatments to examine a longer duration of the observed tRNS effect.

Our results also highlight the importance of combining cognitive training with tRNS. It was suggested that tRNS alone does not yield a lasting behavioral effect (Cappelletti *et al*., 2013; Cohen Kadosh, 2015). Our approach is likely to require that brain stimulation be combined with cognitive training in order to induce lasting effects. By contrast, some of the previous studies are based on stimulating the brain at rest, or applying brain stimulation during sleep (McGough *et al*., 2019). The requirement of active cognitive engagement vs. passive involvement of the patient is important, and caregivers/clinicians will have to consider these options given the commitment constraints of the caregiver but at the same time the potential to induce lasting effects. In addition, current neurostimulation studies do not take into account the suitability of the type of intervention as a function of ADHD subtype. Future studies that will be built on successful protocols could pursue such important directions with a larger sample size that is necessary to address such possibility.

In addition, in the present study we chose to compare the effect of tRNS vs. tDCS, rather than vs. sham stimulation. In our view, such an approach is more rigorous as it compares the effect of two stimulation protocols that at the theoretical and the empirical level had an a priori likelihood of leading to successful treatment. Therefore, the obtained results are expected to be stronger when compared to sham stimulation. However, we would like to acknowledge a potential criticism that tDCS, in contrast to sham stimulation, might yield impairment, rather than improvement. While a future study that includes a sham group is needed to exclude this possibility with great confidence, the criticism is likely unfounded given the accumulated evidence that suggests a beneficial effect of tDCS on ADHD neuropsychological deficits under the montage we used (Munz *et al*., 2015; Bandeira *et al*., 2016; Nejati *et al*., 2017; Allenby *et al*., 2018; Soltaninejad *et al*., 2019). Future research that compares different approaches to intervention, and the mechanisms these interventions are acting on, will advance our understanding of and decision-making on the most promising approaches to ADHD treatment.

Compared to the mechanisms involved in other brain stimulation methods such as transcranial magnetic stimulation, tDCS, or transcranial alternating current stimulation, the neurocognitive mechanisms in tRNS are less known (Chaieb *et al*., 2015; Fertonani and Miniussi, 2017). The most prevalent explanation for tRNS is stochastic resonance. Stochastic resonance describes the phenomenon of introducing an appropriate level of random noise to enhance the output of subthreshold signals. With respect to tRNS, it suggests that the application of weak electric currents amounts to an introduction of neural noise (Fertonani and Miniussi, 2017). Information processing at the neuronal level is sensitive to stochastic resonance (McDonnell and Ward, 2011). tRNS at different intensities over the visual cortex has been shown to lead to behavioural changes in a manner that corresponds to an inverted-U function, a characteristic of stochastic resonance (van der Groen and Wenderoth, 2016).

According to this framework the effect of tRNS in the present study might be attributed to amplifying underactive basal ganglia-thalamo-cortical circuits that has been associated with ADHD (Aron *et al*., 2004; Christakou *et al*., 2004; Norman *et al*., 2016). In this scenario, targeting the prefrontal cortex impact the dorsal neo-striatum via excitatory glutaminergic cells, the basal ganglia to the dorsomedial thalamus via inhibitory projections, and the thalamus back to the prefrontal cortex via excitatory projections (Castellanos *et al*., 2002).

A second mechanistic explanation for the tRNS in the present study is coming from a combined electroencephalography-tRNS study that found that the behavioural improvements of tRNS above the dlPFC vs. sham tRNS are associated with alterations in amplitude of attention and preparatory markers. Those results suggest that the enhancement effect of tRNS when applied above the dlPFC acts by effecting general attentional mechanisms during cognitive training (Sheffield *et al*., 2020). However, it is important to highlight that the abovementioned study included healthy young adults.

However, another possibility is that tRNS impacted oscillations between 140–220Hz (“ripples”), which are involved in learning and long-term potentiation (Ponomarenko *et al*., 2008; Jadhav *et al*., 2016). Ripple oscillations underlie learning via Hebbian requirements for synaptic modification, and are attributed to the hippocampus as well as the prefrontal cortex (Jadhav *et al*., 2016). In awake animals ripple oscillations appear to represent a direct network correlate of learning behaviour (Ponomarenko *et al*., 2008). If tRNS modulated ripple oscillations and by that causally alter learning and long-term potentiation in ADHD then a more optimal tRNS frequencies would be within 140-220Hz, rather than 100-640Hz as in this study. Namely, while in the non-cognitive domain high-frequency tRNS (101–640Hz) has been suggested to be more beneficial than low-frequency (0.1–100Hz) (Terney *et al*., 2008; Fertonani *et al*., 2011), the prediction of the ripple oscillations hypothesis is that the most optimal parameters will appear in the range of 140–220Hz (Ponomarenko *et al*., 2008; Jadhav *et al*., 2016). It might be that transient tRNS effect, as was found in other studies, might be due to other mechanisms such as stochastic resonance, while the neuroplasticity effects could be due to ripple oscillations.

In this regard, a recent study in mice that aimed to advance the understanding of the effect of tRNS on the developing brain has revealed that delivery of identical tRNS current density and duration per day over multiple sessions (in this case 9 sessions, twice a week) to the prefrontal cortex reduces glutamic acid decarboxylase 65/67 but not vesicular glutamate transporter 1. This effect was maximal in the location immediately beneath the electrode but not in a deeper location (Cohen Kadosh *et al*., 2019). Such findings support our suggestion that tRNS impacts neuroplastic mechanisms and, at least in mice, involves the GABAergic system. Further work in humans and animals could shed further light on the mechanisms involved, and on how tRNS can ameliorate ADHD symptoms and potentially other clinical conditions.

## Data Availability

The data is available upon request from the authors.

## Acknowledgements

We thank Prof. Katya Rubia for initiating the idea to use the ACTIVATE^™^ training program with brain stimulation. We also thank Snir Barzilay, Noam Galon, Yehudit Fox, Romy Goldfus and Noa Ariely for their help with data collection, organization and psychological evaluations. The manuscript has been posted on a preprint server (medRxiv).

## Supplemental Information

**Table S1.** Beta Weights (Standardized) of the Regression Model with Post-treatment MOXO Attention Score as the Outcome Measure.

| | $\beta$ | Std.Error | DF | t-value | p-value |
| --- | --- | --- | --- | --- | --- |
| (Intercept) | -0.1335 | 0.0917 | 51 | -1.4563 | 0.1514 |
| MOXO Attention (Baseline) | 0.0903 | 0.0381 | 51 | 2.3684 | 0.0217 |
| Stimulation | -0.0088 | 0.0258 | 51 | -0.3431 | 0.7329 |
| Time | 0.0344 | 0.0257 | 51 | 1.3357 | 0.1875 |

**Table S2.** Beta Weights (Standardized) of the Regression Model with Post-treatment MOXO Impulsivity Score as the Outcome Measure.

| | $\beta$ | Std.Error | DF | t-value | p-value |
| --- | --- | --- | --- | --- | --- |
| (Intercept) | -0.8218 | 0.0305 | 51 | -26.936 | <0.0001 |
| MOXO Impulsivity (Baseline) | -0.0631 | 0.0273 | 51 | -2.306 | 0.0252 |
| Stimulation | -0.0075 | 0.0241 | 51 | -0.3136 | 0.7551 |
| Time | -0.0043 | 0.0238 | 51 | -0.1833 | 0.8553 |

**Table S3.** Beta Weights (Standardized) of the Regression Model with Post-treatment MOXO Hyperactivity Score as the Outcome Measure.

| | $\beta$ | Std.Error | DF | t-value | p-value |
| --- | --- | --- | --- | --- | --- |
| (Intercept) | -0.1453 | 0.0393 | 51 | -3.692 | 0.0005 |
| MOXO Hyperactivity (Baseline) | -0.1414 | 0.0337 | 51 | -4.194 | 0.0001 |
| Stimulation | -0.0217 | 0.0279 | 51 | -0.777 | 0.4404 |
| Time | 0.0154 | 0.0281 | 51 | 0.549 | 0.5852 |

**Table S4.**
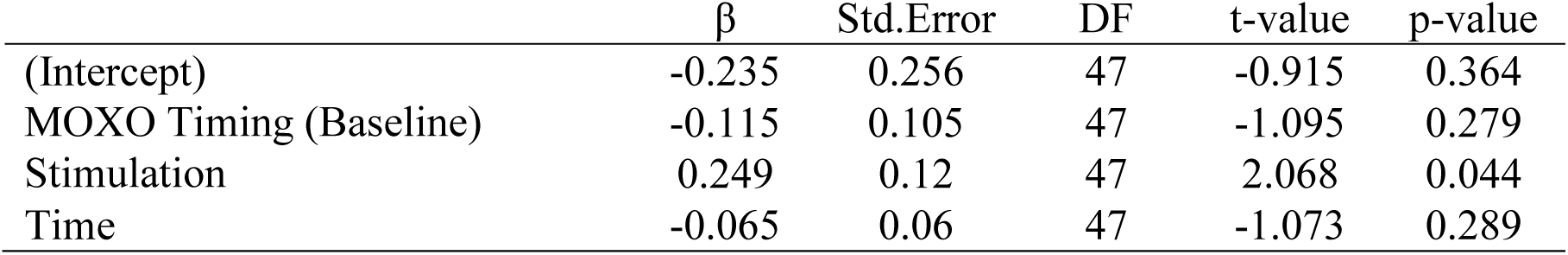
Beta Weights (Standardized) of the Regression Model with Post-treatment MOXO Timing Score as the Outcome Measure.

| | $\beta$ | Std.Error | DF | t-value | p-value |
| --- | --- | --- | --- | --- | --- |
| (Intercept) | -0.235 | 0.256 | 47 | -0.915 | 0.364 |
| MOXO Timing (Baseline) | -0.115 | 0.105 | 47 | -1.095 | 0.279 |
| Stimulation | 0.249 | 0.12 | 47 | 2.068 | 0.044 |
| Time | -0.065 | 0.06 | 47 | -1.073 | 0.289 |

**Table S5.** Beta Weights (Standardized) of the Regression Model with Post-treatment CGI-S Score as the Outcome Measure.

|  | B | Std.Error | DF | t-value | p-value |
| --- | --- | --- | --- | --- | --- |
| (Intercept) | -0.333 | 0.113 | 53 | -2.955 | 0.005 |
| Stimulation | -0.067 | 0.09 | 53 | -0.753 | 0.455 |
| CGI-S (Baseline) | 0.082 | 0.093 | 53 | 0.881 | 0.383 |
| Time | -0.052 | 0.089 | 53 | -0.588 | 0.559 |

**Table S6.** Beta Weights (Standardized) of the Regression Model with Post-treatment Forward Digit Span Score as the Outcome Measure.

| | $\beta$ | Std.Error | DF | t-value | p-value |
| --- | --- | --- | --- | --- | --- |
| (Intercept) | -0.038 | 0.167 | 51 | -0.225 | 0.823 |
| Stimulation | 0.039 | 0.159 | 51 | 0.242 | 0.81 |
| Forward digit span (Baseline) | 0.381 | 0.114 | 51 | 3.328 | 0.002 |
| Time | 0.006 | 0.079 | 51 | 0.698 | 0.945 |

## A description of the Computerized EF training (ACTIVATE^™^)

The computerized training used in the study (ACTIVATE^™^ by c8 sciences, https://www.c8sciences.com/) is comprised of four EF training games, collectively targeting the EF functions of sustained attention, response inhibition, spatial working memory, cognitive flexibility, switching, and divided attention. The code for the games was created with the text editor Sublime (https://www.sublimetext.com/) and artwork created in Photoshop (http://www.photoshop.com/).

Training is delivered on tablets. Each participant logs into the training using a unique, password-protected login. Once the game starts, a pirate island theme is presented, and the child is asked to choose between four different game options. Two of the games (Treasure trunk and Magic lens) mainly target response inhibition, while two others (Monkey trouble and Grub Ahoy) mainly target working memory. The instructions were presented both aurally and visually on the screen at the beginning of each game. The initial sessions of each game are short and easy to master, and the difficulty level was adjusted up or down every 10-15 seconds, according to the individual abilities of each player. In addition, when the child makes an error, the program coaches him/her until he/she successfully corrects the error and moves on to the next level. Auditory and/or visual feedback were given after every trial.

Below we provide details on the four games included in our trial:

A. **Treasure Trunk**. This task trains basic sustained attention abilities at its initial levels, but systematically adds discriminant attention, response inhibition, cognitive flexibility, working memory and multiple simultaneous attention to constitute a general executive function training. In this exercise, children begin by using the mouse to track a moving light and click on it when it turns into a red jewel. With correct responses, the light moves faster, with mistakes it slows down. As the exercise continues, more aspects of executive function are added. Blue jewels appear that should not be clicked, adding discriminant attention and response inhibition. Next, the target switches randomly between red and blue, increasing response inhibition demands and adding cognitive flexibility. Working memory is introduced by showing half-jewels and instructing children to only click on one that is the same color as the one before in order to create a full jewel. In addition to the primary focus on executive functions, this training also requires visual-spatial processing and hand-eye coordination. All levels repeat with two and three moving lights on the screen.
B. **The Magic Lens**. This exercise targets sustained attention, cognitive flexibility, response inhibition and working memory. In this exercise, the child must first find the monkeys hidden in the pirates’ crates. When the magic lens reveals a monkey, the child should click the monkey to free it from the crate. The lens moves faster as the child makes more correct responses, and slows down when the child makes a mistake. In higher levels of difficulty, the child needs to search for other objects in the crates – first pirate monkeys and, later, boots, which is more difficult. The child uses clues at the top of the screen to know what rules they are to follow; which kind of monkey/boot he should free.
C. **Monkey Trouble**. In this exercise, the child sees a pattern of objects appear on screen for a short period of time. When the pattern disappears, the child has to recreate it by clicking on the specific objects in the correct order. The pattern that the child needs to memorise increases with correct answers, and decreases if the child makes a mistake. As the levels of the exercise increase, the child will need to be able to match the correct pattern in reverse.
D. **Grub Ahoy!** In this exercise, the child has to remember the order in which a group of pirates seated on the beach raise their hands to request dinner, or the places in a campsite visited by a playful monkey. The child needs to click on the characters in the appropriate order. The number of locations to be remembered begins with two and can increase with success and decrease with mistakes. Some levels require the child to respond in reverse order.

## Abbreviations

ADHD: Attention deficit hyperactivity disorder
ADHD-RS: ADHD rating scale
CGI-S: Clinical Global Impression – Severity
CI: confidence intervals
tES: transcranial electrical stimulation
tDCS: transcranial direct current stimulation
tRNS: transcranial random noise stimulation
dlPFC: dorsolateral prefrontal cortex
EF: executive function

